# HiFi long-read genomes for difficult-to-detect clinically relevant variants

**DOI:** 10.1101/2024.09.17.24313798

**Authors:** Wolfram Höps, Marjan M. Weiss, Ronny Derks, Jordi Corominas Galbany, Amber den Ouden, Simone van den Heuvel, Raoul Timmermans, Jos Smits, Tom Mokveld, Egor Dolzhenko, Xiao Chen, Arthur van den Wijngaard, Michael A. Eberle, Helger G. Yntema, Alexander Hoischen, Christian Gilissen, Lisenka E.L.M. Vissers

## Abstract

Clinical short-read exome and genome sequencing approaches have positively impacted diagnostic testing for rare diseases. Yet, technical limitations associated with short reads challenge their use for detection of disease-associated variation in complex regions of the genome. Long-read sequencing (LRS) technologies may overcome these challenges, potentially qualifying as a first-tier test for all rare diseases. To test this hypothesis, we performed LRS (30x HiFi genomes) for 100 samples with 145 known clinically relevant germline variants that are challenging to detect using short-read sequencing and necessitate a broad range of complementary test modalities in diagnostic laboratories.

We show that relevant variant callers readily re-identify the majority of variants (120/145, 83%), including ∼90% of structural variants, SNVs/InDels in homologous sequences and expansions of short tandem repeats. Another 10% (n=14) was visually apparent in the data but not automatically detected. Our analyses also identified systematic challenges for the remaining 7% (n=11) of variants such as the detection of AG-rich repeat expansions. Titration analysis showed that 89% of all automatically called variants could also be identified using 15-fold coverage.

Thus, long-read genomes identified 93% of pathogenic variants that are most challenging to detect using short-read technologies. Even with reduced coverage, the vast majority of variants remained detectable, possibly enhancing cost-effective diagnostic implementation. Most importantly, we show the potential to use a single technology to accurately identify all types of clinically relevant variants.

---

The >7,000 rare diseases (RD) known to date collectively present a common healthcare issue. More than 70% of RD are genetic in origin, and their molecular genetic diagnosis is important for patients and families (Nguengang Wakap et al. 2020). Comprehensive diagnostics of rare genetic disease requires a complex mix of diverse testing modalities. Many diagnostic laboratories still apply traditional approaches such as karyotyping, FISH, genomic microarrays, Southern blotting, MLPA, and Sanger sequencing, leading to complex, long-lasting and often expensive testing cascades. Over the last decade, next generation sequencing (NGS), particularly exome and genome sequencing (ES and GS, respectively) have emerged as more generic clinical tests (Wojcik et al. 2024; Turro et al. 2020). While GS represents the most successful first tier test to date, a recent systematic study showed that short read sequencing-based GS would not be suited to replace all other diagnostic approaches for up to 25% of all patients referred for testing (Schobers et al. 2024). For this group of patients, technical limitations associated with short read technologies prevent the robust identification of certain clinical variant types. These include, but are not limited to, short tandem repeat (STR) expansions, complex structural rearrangements (such as translocation, complex structural variations (SVs), Mobile Element Insertions (MEI)) and variants in segmental duplications, genes with homologies and pseudogenes. Other factors that make the detection of specific variants with short-read whole genome sequencing (SR-WGS) challenging include extreme GC content and homopolymer stretches. Moreover, certain readouts such as methylation were never possible and required separate test modalities. Long-read sequencing (LRS) may, however, overcome many of these challenges. Emerging LRS technologies include PacBio HiFi LRS, Oxford Nanopore Technologies (ONT) nanopore-based sequencing, and synthetic long-read technologies such as iCLR (Gorzynski et al. 2024). These technologies have matured to enable population-level sequencing (Schloissnig et al. 2024; Gustafson et al. 2024; Beyter et al. 2021) and have led to discoveries in rare disease research (Mantere, Kersten, and Hoischen 2019). A systematic assessment of their application in clinical diagnosis has, however, yet to be performed.

We assessed the clinical utility of HiFi long-read genome sequencing (LRS) by testing 100 samples with 145 known clinically relevant variants, specifically enriched for variants that are challenging or impossible to identify by SR-WGS (**Figure 1A-B** and **Table S1)**. Of note, 42 of the 100 samples, containing 70 variants, were previously included in a clinical utility study using 30x short-read genomes (Schobers et al. 2024). This earlier study demonstrated the challenges of SR-WGS variant detection as only 29 of the 70 variants could be identified using automated variant calling algorithms. Automated detection was challenging because of the type of variants (e.g. STRs and SVs) or their location within the genome (e.g. homologous regions). The remaining 58 samples were chosen to contain similarly difficult variant types or variants located in similarly difficult regions of the genome. Hence, the total series included 25 samples with STR expansions across 14 different loci, 36 samples with variants in 16 different regions characterized by homology and/or pseudogenes, and 24 samples with (complex) structural events. The remaining 15 samples included a variety of other clinically relevant variants such as imprinted loci and mtDNA variation.

**Figure 1.**
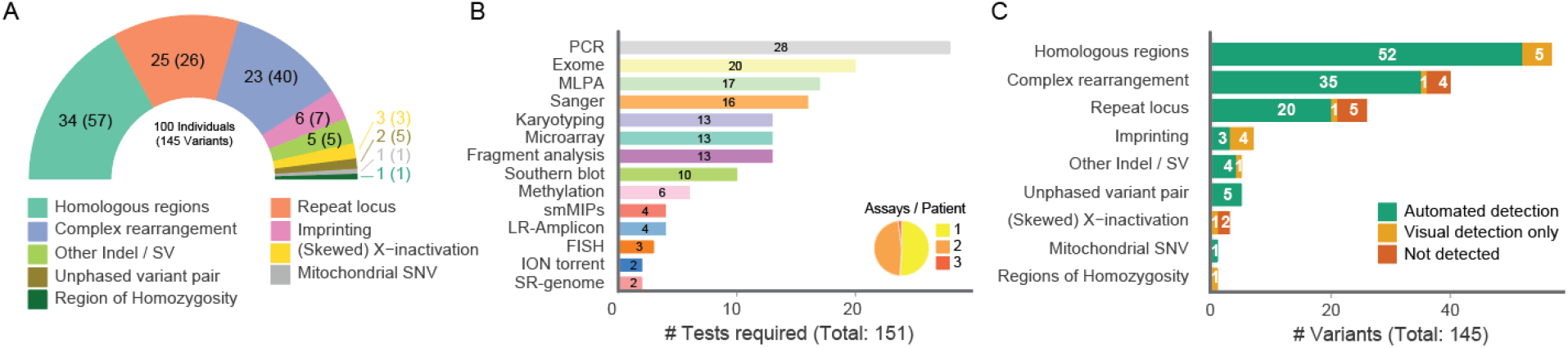
Samples, variants and LRS-based recovery. **(A)** Pie chart depicting the cohort composition by variant type for all 145 variants. The number of samples is indicated within parentheses. **(B)** Different test modalities (y-axis) that were used in a diagnostic laboratory to identify all 145 clinically relevant variants in the 100 selected samples (x-axis). The number of assays required per patient is shown in an inlay pie chart **(C)** Sensitivity of LRS by automated variant detection and visual inspection for all 145 variants from 100 analyzed samples (x-axis), stratified by *a-priori* known disease-associated variant types (y-axis). LRS-based detection rates are indicated in green (detected by a variant caller, 83% (n = 120)), orange (detection by visual read inspection, 10% (n = 14)) and red (undetected variant, 7% (n = 11)).

Here, we used a single SMRT cell on a PacBio Revio system for each sample, expected to generate ±30x coverage. All samples were processed in the same fashion, according to the manufacturer’s instructions (PacBio, Menlo Park, CA, USA). In brief, 7 μg high molecular weight gDNA was sheared on Megaruptor 3 (Diagenode, Liège, Belgium) to a target size of 15-18 kb, libraries were prepared with SMRTbell prep kit 3.0 (PacBio, Menlo Park, CA, USA), size-selected□>10 kb on the BluePippin (Sage Science, Beverly, MA, USA), and sequenced for 24 h on the Revio system (ICS 12.0.4). Samples were then analysed using a bioinformatics pipeline that incorporates a variety of PacBio software tools for long read sequencing (LRS) data analysis. Alignment (pbmm2 v1.10.0) of High Fidelity (HiFi) reads was performed against the GRCh38 reference genome, while generating haplotags by performing *de novo* assembly with Hifiasm (v.0.15.3). Structural variants (PBSV v2.9.0) and Small variants (DeepVariant v1.5.0) were called with phasing information (HiPhase v0.10.1) and annotated using publicly available databases. An additional analysis for copy number variants (CNV) based on read depth was performed using HiFiCNV (v0.1.6). Short tandem repeats were called (TRGT v0.4.0), visualized (TRVZ v0.4.0) and annotated using an inhouse pipeline. Specific variant calls for paralogs and pseudogenes are called using Paraphase (v.2.2.3). Methylation calls were generated using pb-cpg-tools (v.2.3.1). When the expected variant was not detected by the respective software, the sequencing data were visually inspected using the Integrated Genomics Viewer (IGV, v2.16.2) for region(s) containing the variant(s), taking into account a suitable genomic window. The mode of visual confirmation depended on the variant type and variants were considered visually confirmed when sequencing reads showed a clear deviant pattern in read mapping, methylation profiles or variant allele frequencies.

Across all 100 samples, we obtained a median output of 94.0 Gb of data, achieving a median genome-wide coverage of 29.7x, with an average read length of 15.35 kb (**Table S2**). Of the 145 variants analyzed, 120 (83%) were detected fully automatically by the relevant variant calling software (**Figure S1**). This included, among others, 61 SVs, 20 STR expansions, and 40 SNV/indels the majority of which affected loci with homologous sequences (**Table S1, Figure 1C**). The 20 repeat expansions ranged from 16 to >150 additional repeat units in 12 different genes. In some cases, HiFi genomes also provided additional molecular insights that were not obtained from the traditional clinical tests. For instance, targeted *de novo* assembly by HiFiasm (version 0.15.3) and visualization by NAHRwhals v1.4 (Höps et al. 2023) allowed us to resolve the structure of a complex genomic rearrangement encompassing the *CEP85L* and *MCM9* genes (**Figure 2F**). In addition, in one case, two additional smaller duplications were identified adjacent to the known pathogenic variant; and in another case involving the *OPN1LW*/*OPN1MW* gene cluster (P11-F11), the sequence context of the second *OPN1MW* copy was revisited now that the entire copy could be read at base pair level. While similar benefits of SR-WGS have been reported from direct comparisons of short-read genomes to exomes (van der Sanden et al. 2023), the ability to analyze even these complex loci of the human genome using LRS provides great promises for clinical care.

**Figure 2.**
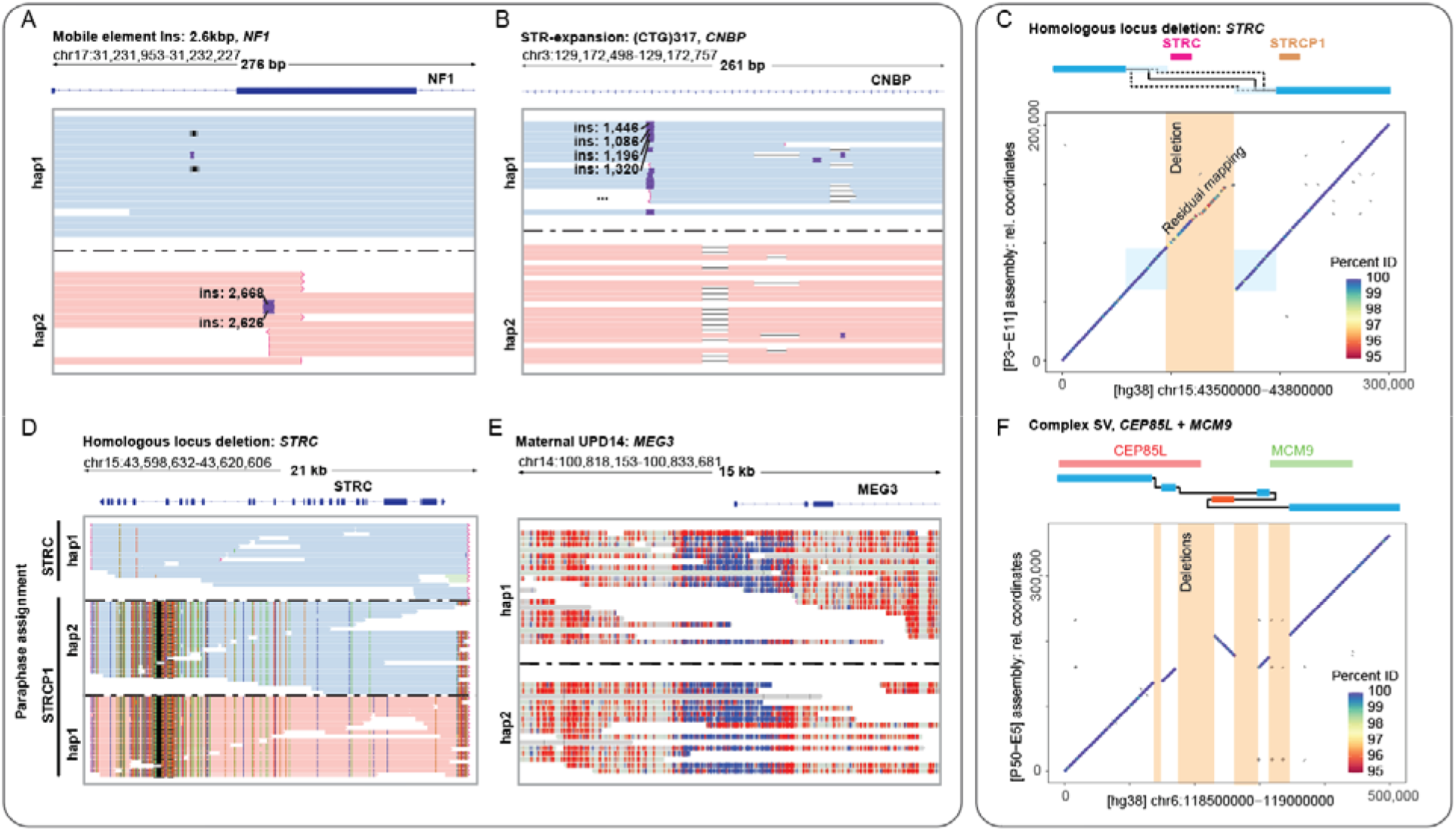
Examples of variants identified in an automated fashion or by visual inspection. **(A-B)** IGV screenshots of long-read sequencing data for specific variants. Reads are colored by phase. (**C**) Visualization of the *de novo* assembly of a deletion of *STRC*, with the pseudogene *STRCP1* intact. Using the mapping quality metric, the breakpoint can be narrowed down to a ∼30kbp window (light blue squares). A schematic view of the genes and assembly mapping is indicated on top, raw dotplot mappings of GRCh38 (x-axis) vs the assembled region (y-axis) are displayed below. (**D)** The same variant from (C) visualized with Paraphase. Reads are grouped by inferred (pseudo)gene identity. Only one haplotype of *STRC* is observed, thus indicating a deletion of the other allele. (**E)** An imprinting defect on the maternal chromosome 14 due to a uniparental heterodisomy. Reads are colored by methylation status, with blue indicating unmethylated CpGs and red methylated CpGs. (**F**) *De novo* assembly of a locus containing a ∼200kbp complex genomic rearrangement.

For an additional 10% (n=14) of variants (**Figure S1**), automated detection failed, requiring either manual inspection of aligned sequencing reads (n=8) or visualization of processed data files in the absence of dedicated callers (n=6; **Figure S2)**. Manual inspection successfully identified variants in the *OPN1LW/OPN1MW* (n=3) and *CFHR* (n=2) gene clusters as well as a complex structural rearrangement involving complex/repetitive regions, one of the STRs (in *CNBP*), and a MEI inserted in *NF1*. Here, too, molecular insights were further enhanced by *de novo* assemblies, for instance discriminating between the deletion of the *STRC* gene and its *STRCP1* pseudogene copy in one of the samples (P3-E11; **Figure 2C, Supplementary Methods**). We expect that further improvements in structural variant detection based on *de novo* assembly should allow for automatic detection of these variants in the future. Nonetheless, as obtaining contiguous genome-wide *de novo* assemblies is still challenging, it may be especially beneficial for long-read technologies to use the most accurate human reference genome for mapping purposes, such as the one presented by the telomere-to-telomere (T2T) consortium (Aganezov et al. 2022). Indeed, in one of the samples (P50-E2), an unbalanced translocation involving chromosomes 13 and Y, the variants could only be fully resolved when mapping to the T2T reference instead of GRCh38.

For six variants, evaluation of detection could only be performed after additional computational analyses of the output files generated by the standardized variant callers included in our pipeline (**Methods**). Three of these variants concerned regions of homozygosity (ROH) for which no appropriate calling algorithm was available (**Figure S2**). For these, we visualized homozygosity by plotting variant allele frequencies of SNPs within the region (e.g. B-allele frequency plots; **Methods**), allowing the detection of each of them. We anticipate that methods for automatically detecting ROH and uniparental disomies (UPD, (Yauy et al. 2020)) from SR-WGS can be readily adapted for long-read data, enabling automated detection of these variants. The other three variants concerned various percentages of skewed X-inactivation (P5-H5 80/20%; P50-A6, 90/10% and P50-E8, 100/0%). To detect these variants, we devised a method to calculate the ratio of methylation per allele in all female samples (**Methods**). Only one of three showed a clear deviation in the methylation pattern confirming skewing (P50-E8; **Figure S2**), whereas the other two could not be discriminated from the other 49 tested female samples (**Figure S3**). This suggests that LRS either lacks the sensitivity to detect skewing below 80% (at 30x coverage), or that the standard-of-care method to detect skewed X-inactivation involving methylation status of a single locus (the *AR* gene) may not be predictive for the methylation status of the entire X-chromosome. However, targeted interpretation of the *AR* locus in LRS data of these two samples also did not suggest skewing. Notably, two other samples (P2-E4, P1-C1) identified by our analysis as potentially having skewed X-inactivation had pathogenic structural variants on the X-chromosome. It is to be expected that new methods for the automated calling of methylation defects from LRS data will further enhance the utility of LRS in a clinical setting.

Of all 145 variants, 11 (7%) could not be detected in HiFi genomes, neither by automated callers or by visual inspection, nor by additional computational analyses. We investigated whether a common factor could explain these detection failures. Two of the variants were related to the unverifiable skewed X-inactivation described above. Five variants (3%) involved GA-based repeat expansions, including (GAA)_n_ in *FXN* and (AAGGG)_n_, (AAAGG)_n_, or (ACAGG)_n_ in *RFC1*, suggesting a systematic issue (**Table S1**). For these variants, HiFi sequencing suffered from reduced quality, resulting in fewer high quality reads available for HiFi read generation, leading to insufficient sequencing coverage in these regions for variant calling. This reduced quality for GA-repeat regions may result from the formation of non-B DNA conformations, hampering the DNA polymerase and reducing read length and quality (Mellor, Perez, and Sale 2022). Based on this assumption, we hypothesized that if non-HiFi quality reads could be ‘rescued’, additional coverage could be added for these regions, potentially allowing for better calling of repeat lengths. Indeed, by manually adding “rejected” low quality reads to the HiFi data, we increased the coverage for *FXN*, enabling the automatic detection of the pathogenic repeat (**Figure S4, Table S3**). However, this approach did not work for *RFC1* repeats where coverage remained too low for detecting the expansion in all four cases. Notably, sufficient coverage is observed at these repeat regions for wildtype alleles, with reduced coverage only apparent in cases of repeat expansions. This observation supports our hypothesis of allelic drop out due to technical difficulties as a consequence of the repeat expansion. Hence, detecting reduced coverage at these repeat loci could serve as an indicator of a potential repeat expansion, warranting further follow-up. This proxy detection method of repeat expansions might be useful when HiFi genomes are used as a first-tier test. Alternatively, given the clinical recognizable phenotypes associated with most GA-based repeat expansions (e.g. rare neurological movement disorders), a targeted approach, such as the PureTarget technology, may also be considered as the increase in read depth for those loci would potentially compensate for the loss of high-quality reads, allowing automated calling.

For the remaining four variants that could not be detected, detection was hampered by the fact that each variant was associated with breakpoints in highly repetitive regions (P4-C4 and P3-F6) or segmental duplications >50kb in size (P1-C1 and P9-B1). For P4-C4, a translocation between the acrocentric p-arm of chromosome 22 and the repeat-rich regions of the Y-chromosome (P4-C4), and a P3-F6, involving a Robertsonian translocation affecting the acrocentric p-arms of chromosomes 13 and 14, we attempted re-alignment using the T2T reference genome but this did not recover the variants. P1-C1 and P9-B1 concerned unbalanced translocation events whose breakpoints fell inside segmental duplications, leading to their incorrect classification as simple copy number variants. Detecting these four variants may require further algorithmic development or the use of long range DNA information that can span regions >50kb, as recently demonstrated by others (Guarracino et al. 2023).

We next focused our attention on a technical evaluation of the role of sequencing coverage. Several studies have assessed minimum coverage thresholds for LRS (Noyes et al. 2022; Lee, Kim, and Lee 2023). However, these studies either relied on gold standards of unselected variants that were also detectable with SR-WGS, were based on non-human models (Lee, Kim, and Lee 2023) or alternatively focused on one specific class of variants (Noyes et al. 2022). Given our unique heterogeneous set of validated variants, we saw an opportunity to estimate the sensitivity for detection in relation to depth of coverage. To this end, we implemented an *in-silico* downsampling experiment (**Supplementary Methods**). Three samples (P13-G4, P50-G5, P50-H7) with an initial coverage of less than 20-fold were excluded from this analysis. For the remaining samples, BAM files were subsampled to target coverages of 10X, 15X and 20X using the “*samtools view -b”* command (samtools version 1.11). This process was repeated 10 times for each target coverage, resulting in a total of 30 subsampled readsets per sample, or 10 per target coverage. Downstream processing and variant calling were performed analogously to the original datasets, resulting in independent variant callsets for each permutation. In the original dataset, these 97 samples contained 117 biological variants that were detectable by automated callers. Due to cross-calling between different tools (e.g., hifiCNV and pbsv both calling certain CNVs; **Table S1**), those 117 variants were represented by 171 individual variant ‘calls’, which were tested for recall separately (**Table S4**).

We found a reduced discovery rate from initially 171 variants identified at ∼30x, with median detection rates dropping to 162 (96.2%), 151 (89.3%) and 129 (76.3%) for 20x, 15x and 10x coverages, respectively (**Figure 3, Table S4**). Further stratification by variant type and caller revealed the sharpest decline in repeat-associated CNVs and SNVs (recalled by Paraphase) (−18.8% and -14.3% in 15x vs 30x), as well as SVs (−11.1%), CNVs (−8.3%) and SNV/Indels in homologous regions (−10.4%). STRs (−2.8%) and SNV outside of homologous regions (−0%) were less or not at all affected (**Table S4**). We also noted that the reduced sensitivity for SNVs at 20x coverage was entirely driven by those within homologous regions, whereas sensitivity for SNVs outside these regions only diminished from 15x coverage and lower. Our results suggest that 30x or higher coverage may be needed to capture difficult-to-detect variants and harness the full potential of LRS sequencing. However, we also observe that at 15x the primary impact is on the sensitivity for detecting variants in homologous regions. In contrast, the sensitivity of SNVs, CNVs, SVs and STRs in other genomic regions is reduced only by about 10%. Depending on circumstances, the ability to sequence more samples at the same costs may outweigh this reduced sensitivity.

**Figure 3.**
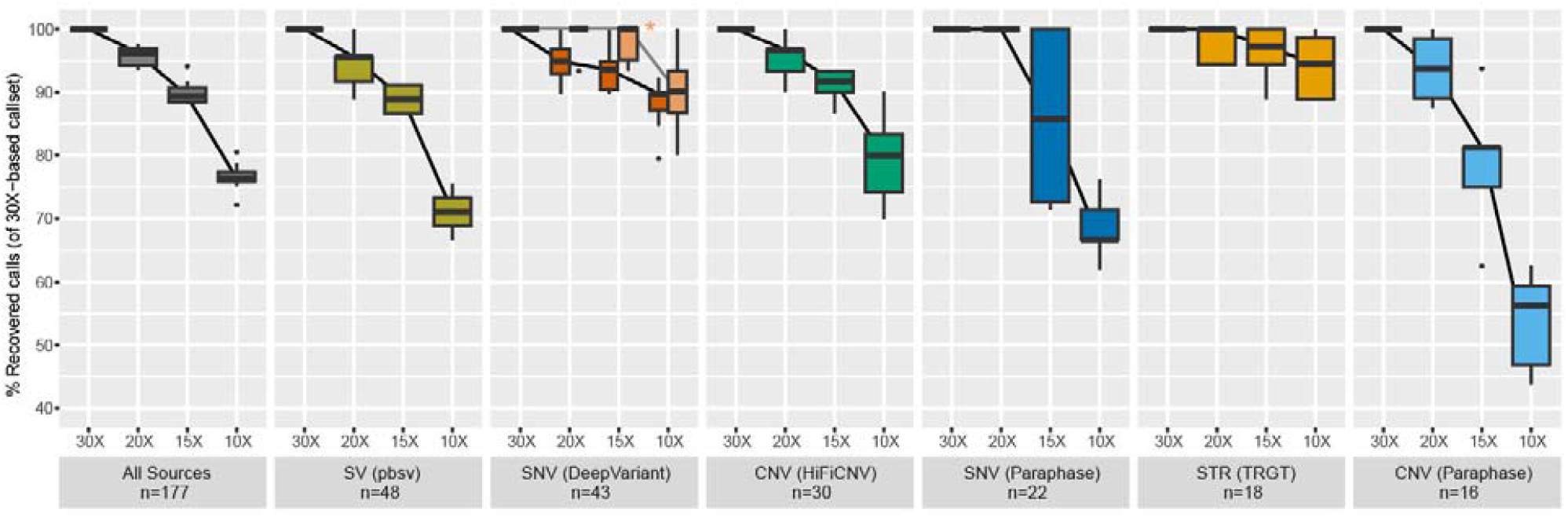
Variant recall in titration experiments. Results of automated variant detection per variant and calling tool for different genome-wide coverage levels (10x, 15x, 20x and 30x) based on the initial 171 calls (**Table S4**). Boxplots are based on 10 random selections of different reads from the original 30x coverage sample. For SNVs we distinguished between all SNVs (in red; n = 43) and SNVs not overlapping a homologous region (in light orange with asterisk; n = 18).

Overall, our data show that HiFi variant calling methods could potentially lead to an automated detection rate of 93% of variants tested. Nonetheless, it should be noted that the results of this study may be confounded by the initial selection of 100 samples. As we select for difficult-to-detect clinically relevant variants, it is anticipated that future use in unselected clinical cohorts the detection rate may be even higher. Our outcomes are of particular relevance in the context of genetic diagnostic laboratories that are considering short- and long-read approaches for generic first-tier germline testing for rare disease. As mentioned, a subset of 42 samples tested here (70 variants) were previously also used to technically benchmark short-read genome sequencing. This unique setup allows for a direct comparison between short- and long-read sequencing for these difficult-to-detect regions of the genome. In short-read genomes, 29/70 variants (41%) were automatically detected compared to 62/70 variants (89%) in HiFi long-read genomes (**Table S5, S6**). Whereas 21 variants in the short-read data could be recovered by visual inspection of the aligned reads, this approach is only feasible for small genomic loci or a limited set of genes, requiring a strong clinical diagnosis to guide interpretation (Corominas et al. 2022). Along with previous studies that have demonstrated the ability to identify molecular diagnoses in a significant percentage of previously undiagnosed cases (Steyaert et al. 2024; Schmidt et al. 2015), our results suggest that HiFi genomes may be a more attractive first-tier, generic assay for germline testing in rare diseases.

In summary, our study provides detailed insights into the abilities and limitations of LRS to identify the full spectrum of clinically relevant genome variation. Although prospective studies are still needed, our results show that LRS has the potential to be implemented as a first-tier diagnostic workflow for germline testing, potentially replacing all current tests for diagnosing individuals with rare disease.

## Data Availability

All relevant data supporting the conclusions of this study, including anonymized variant calls, are contained in the manuscript. The raw sequencing data are not publicly available due to patient confidentiality, but can be made available upon reasonable request to the authors, in accordance with institutional and ethical guidelines.

## Abbreviations

FISH: Fluorescence In Situ Hybridization
IGV: Integrative Genomics Viewer
NGS: Next Generation Sequencing
LRS: Long read Sequencing
MEI: Mobile Element Insertion
MLPA: Multiplex Ligation Probe Amplification
ONT: Oxford Nanopore Technologies
RD: Rare Disease
ROH: Region of Homozygosity
SNV: Single Nucleotide Variant
SRS: Short read Sequencing
SR-WGS: Short read Whole Genome Sequencing
STR: Short Tandem Repeat
SV: Structural Variant
UPD: Uniparental Disomy
WGS: Whole Genome Sequencing

## Acknowledgments

We thank Dr. R. Blok, T. Hofste, Dr. L. Haer-Wigman, Dr. E. Kamsteeg, Dr. N. de Leeuw, Dr. D. Lugtenberg, Dr. T. Rinne, Dr. A. Simons, Dr. C. Harteveld and R. Smeets for useful discussions. We thank the Klinisch Genetisch Centrum Nijmegen, Radboud Genome Technology Center and the Netherlands X-omics Initiative NWO (project 184.034.019), for technical and financial support. The aims of this study contribute to the Solve-RD project (to AH, CG and LELMV), which has received funding from the European Union’s Horizon 2020 research and innovation program under grant agreement No 779257.

## References

Aganezov, Sergey, Stephanie M. Yan, Daniela C. Soto, Melanie Kirsche, Samantha Zarate, Pavel Avdeyev, Dylan J. Taylor, et al. 2022. “A Complete Reference Genome Improves Analysis of Human Genetic Variation.” Science 376 (6588): eabl3533.

Beyter, Doruk, Helga Ingimundardottir, Asmundur Oddsson, Hannes P. Eggertsson, Eythor Bjornsson, Hakon Jonsson, Bjarni A. Atlason, et al. 2021. “Long-Read Sequencing of 3,622 Icelanders Provides Insight into the Role of Structural Variants in Human Diseases and Other Traits.” Nature Genetics 53 (6): 779–86.

Corominas, Jordi, Sanne P. Smeekens, Marcel R. Nelen, Helger G. Yntema, Erik-Jan Kamsteeg, Rolph Pfundt, and Christian Gilissen. 2022. “Clinical Exome Sequencing-Mistakes and Caveats.” Human Mutation 43 (8): 1041–55.

Gorzynski, John E., Shruti Marwaha, Chloe M. Reuter, Tanner Jensen, Alexis Ferrasse, Archana Raja, Liliana Fernandez, et al. 2024. “Clinical Application of Complete Long Read Genome Sequencing Identifies a 16kb Intragenic Duplication in EHMT1 in a Patient with Suspected Kleefstra Syndrome.” medRxiv, 2024–2003.

Guarracino, Andrea, Silvia Buonaiuto, Leonardo Gomes de Lima, Tamara Potapova, Arang Rhie, Sergey Koren, Boris Rubinstein, et al. 2023. “Recombination between Heterologous Human Acrocentric Chromosomes.” Nature 617 (7960): 335–43.

Gustafson, Jonas A., Sophia B. Gibson, Nikhita Damaraju, Miranda Pg Zalusky, Kendra Hoekzema, David Twesigomwe, Lei Yang, et al. 2024. “Nanopore Sequencing of 1000 Genomes Project Samples to Build a Comprehensive Catalog of Human Genetic Variation.” medRxiv : The Preprint Server for Health Sciences, March. 10.1101/2024.03.05.24303792.

Karolchik, Donna, Angela S. Hinrichs, Terrence S. Furey, Krishna M. Roskin, Charles W. Sugnet, David Haussler, and W. James Kent. 2004. “The UCSC Table Browser Data Retrieval Tool.” Nucleic Acids Research 32 (Database issue): D493–96.

Lee, Hyunji, Jun Kim, and Junho Lee. 2023. “Benchmarking Datasets for Assembly-Based Variant Calling Using High-Fidelity Long Reads.” BMC Genomics 24 (1): 148.

Mantere, Tuomo, Simone Kersten, and Alexander Hoischen. 2019. “Long-Read Sequencing Emerging in Medical Genetics.” Frontiers in Genetics 10 (May):426.

Mellor, Christopher, Consuelo Perez, and Julian E. Sale. 2022. “Creation and Resolution of Non-B-DNA Structural Impediments during Replication.” Critical Reviews in Biochemistry and Molecular Biology 57 (4): 412–42.

Nguengang Wakap, Stéphanie, Deborah M. Lambert, Annie Olry, Charlotte Rodwell, Charlotte Gueydan, Valérie Lanneau, Daniel Murphy, Yann Le Cam, and Ana Rath. 2020. “Estimating Cumulative Point Prevalence of Rare Diseases: Analysis of the Orphanet Database.” European Journal of Human Genetics: EJHG 28 (2): 165–73.

Noyes, Michelle D., William T. Harvey, David Porubsky, Arvis Sulovari, Ruiyang Li, Nicholas R. Rose, Peter A. Audano, et al. 2022. “Familial Long-Read Sequencing Increases Yield of de Novo Mutations.” American Journal of Human Genetics 109 (4): 631–46.

Schloissnig, Siegfried, Samarendra Pani, Bernardo Rodriguez-Martin, Jana Ebler, Carsten Hain, Vasiliki Tsapalou, Arda Söylev, et al. 2024. “Long-Read Sequencing and Structural Variant Characterization in 1,019 Samples from the 1000 Genomes Project.” bioRxiv. 10.1101/2024.04.18.590093.

Schmidt, Ellen M., Ji Zhang, Wei Zhou, Jin Chen, Karen L. Mohlke, Y. Eugene Chen, and Cristen J. Willer. 2015. “GREGOR: Evaluating Global Enrichment of Trait-Associated Variants in Epigenomic Features Using a Systematic, Data-Driven Approach.” Bioinformatics 31 (16): 2601–6.

Schobers, Gaby, Ronny Derks, Amber den Ouden, Hilde Swinkels, Jeroen van Reeuwijk, Ermanno Bosgoed, Dorien Lugtenberg, et al. 2024. “Genome Sequencing as a Generic Diagnostic Strategy for Rare Disease.” Genome Medicine 16 (1): 32.

Steyaert, Wouter, Lydia Sagath, German Demidov, Vicente A. Yépez, Anna Esteve-Codina, Julien Gagneur, Kornelia Ellwanger, et al. 2024. “Unravelling Undiagnosed Rare Disease Cases by HiFi Long-Read Genome Sequencing.” medRxiv : The Preprint Server for Health Sciences, May. 10.1101/2024.05.03.24305331.

Turro, Ernest, William J. Astle, Karyn Megy, Stefan Gräf, Daniel Greene, Olga Shamardina, Hana Lango Allen, et al. 2020. “Whole-Genome Sequencing of Patients with Rare Diseases in a National Health System.” Nature 583 (7814): 96–102.

Wojcik, Monica H., Gabrielle Lemire, Eva Berger, Maha S. Zaki, Mariel Wissmann, Wathone Win, Susan M. White, et al. 2024. “Genome Sequencing for Diagnosing Rare Diseases.” The New England Journal of Medicine 390 (21): 1985–97.

Yauy, Kevin, Nicole de Leeuw, Helger G. Yntema, Rolph Pfundt, and Christian Gilissen. 2020. “Accurate Detection of Clinically Relevant Uniparental Disomy from Exome Sequencing Data.” Genetics in Medicine: Official Journal of the American College of Medical Genetics 22 (4): 803–8.

