## Supplementary Figures and Tables for "HiFi long-read genomes for difficult-to-detect clinically relevant variants"

**Supplementary Materials**

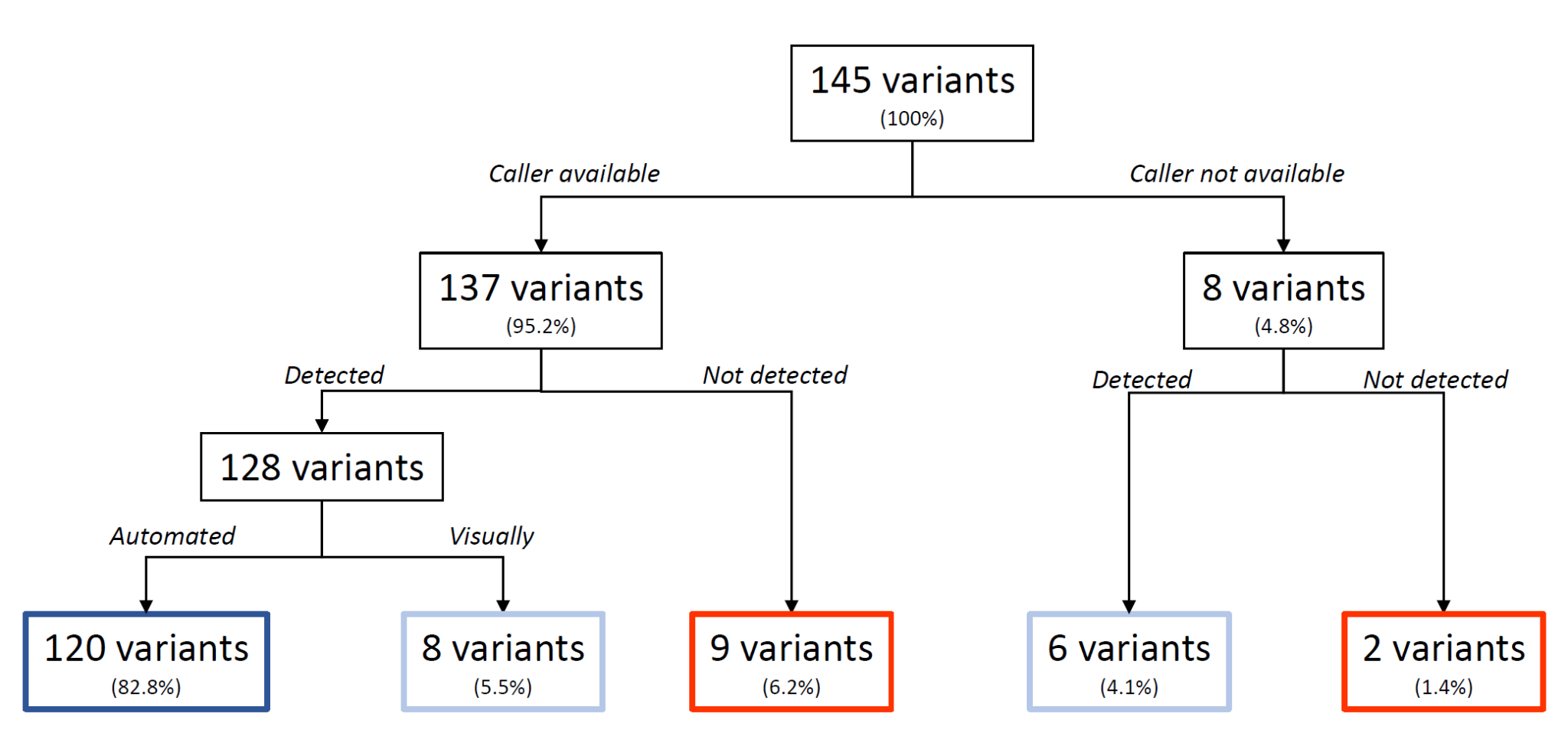

**Figure S1**. Schematic overview of variant detection in 100 samples sequenced at 30x using HiFi long read sequencing. Variants in dark blue boxes were called by standardized callers using automated detection. Variants in the light blue boxes were detected, but required either visual inspection of aligned reads in IGV (for those where callers existed) or alternative methods such as plotting of methylation profiles and/or B-allele frequency plots if no callers were available. The variants in red boxes were not detected. Overall, 83% of variants were detected automatically, another 10% was recovered by additional efforts, and 7% could not be retrieved from 30x HiFi genomes.

**Figure S2**. Examples of IGV screenshots that were used to identify variants through manual inspection.

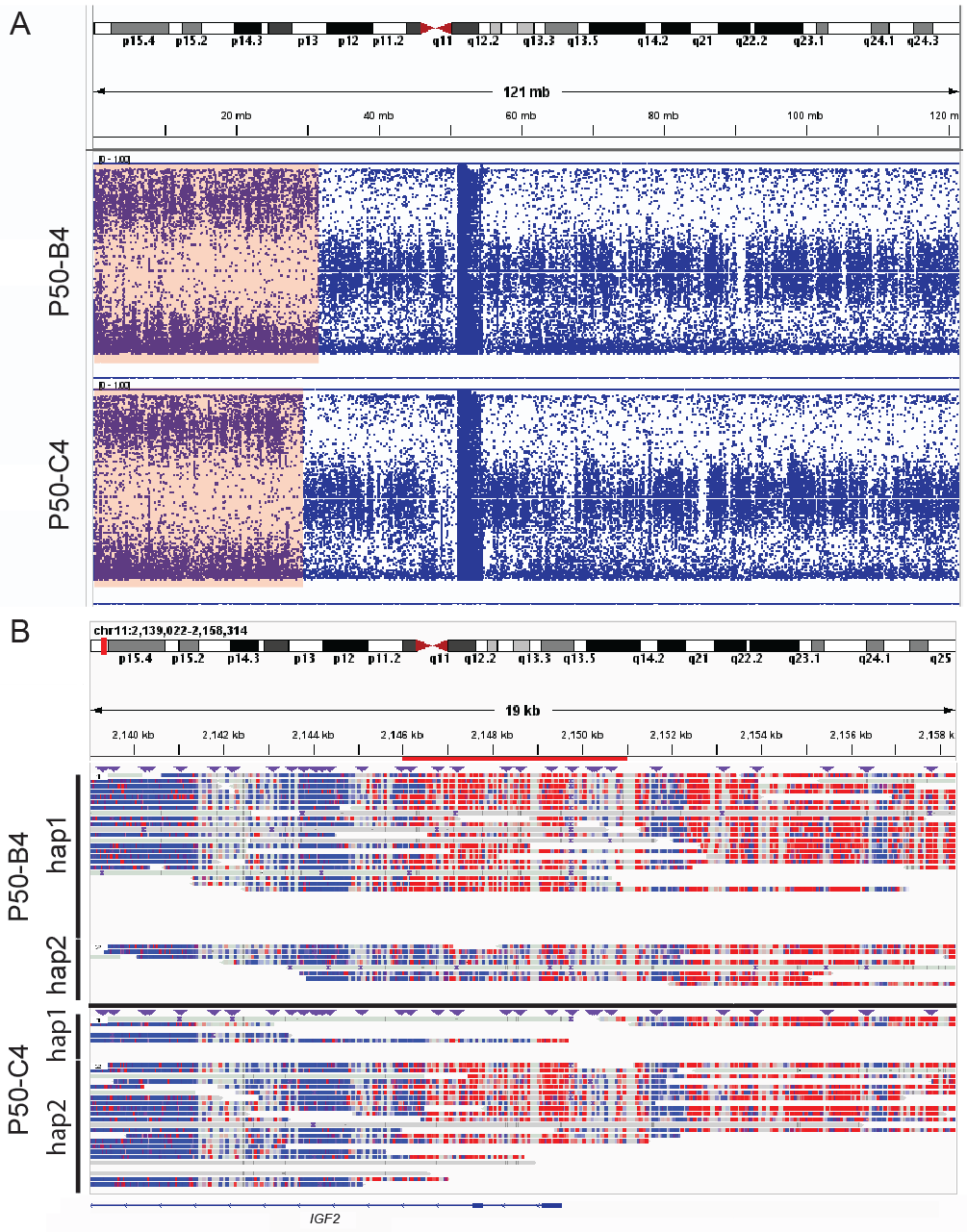

**Figure S2-1.** **Imprinted loci and Regions of Homozygosity. A** IGV screenshot of the Variant Allele Frequency (VAF) of SNPs on chr11. For better visual clarity, only every 5th SNP is plotted. Heterozygous regions (e.g. both q-arms) are expected to show a tri-modal distribution (ref/het/hom) of VAFs, homozygous regions a bi-modal one (ref/alt). The p-arms of both patients show >25 Mb stretches of homozygosity (orange highlights). **B** A zoomed-in view of the imprinted region in the *IGF2* gene. Both samples are mosaic carriers for the ROH, with a fraction of cells still carrying the ‘lost’ haplotype (hap2 in P50-B4 and hap1 in P50-C4). The imprinted region (marked with a red highlight at the top) can be seen to be differentially methylated between the two haplotypes.

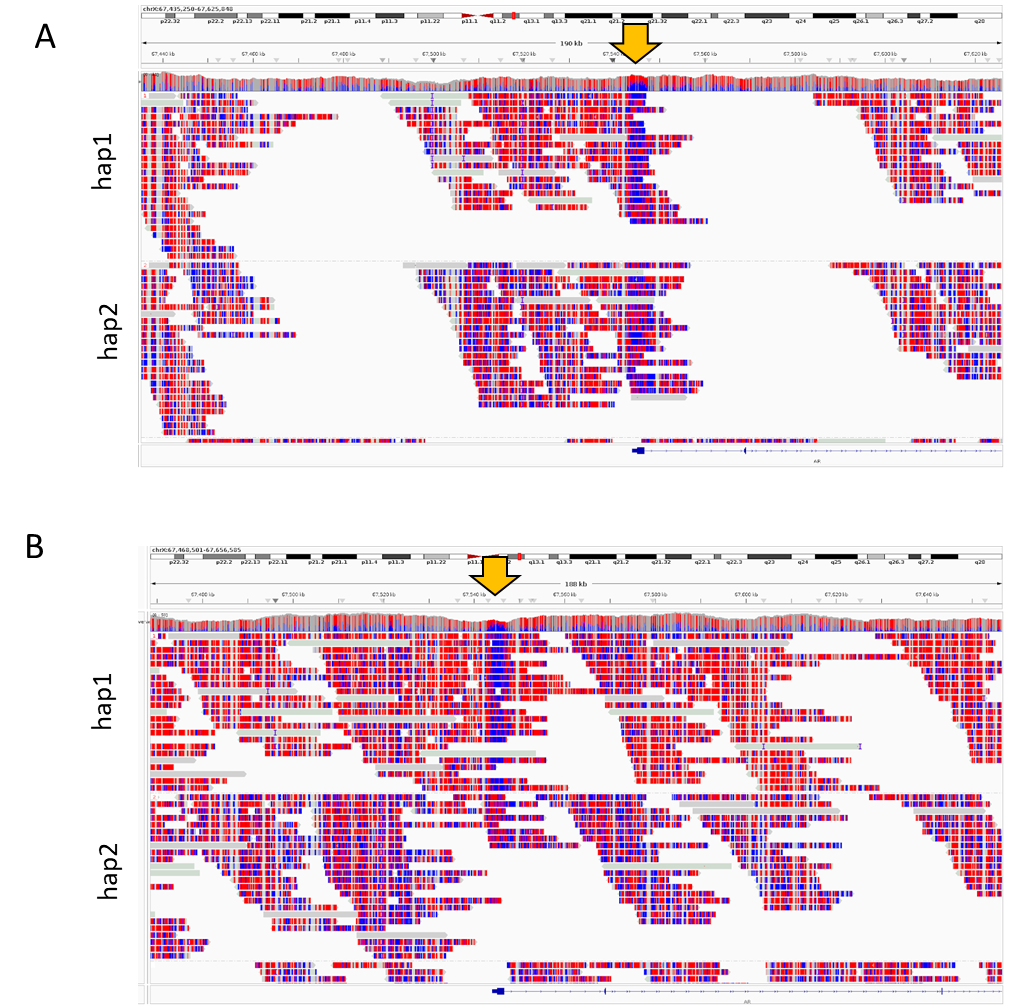

**Figure S2-2: X-inactivation. A** Methylation by long read sequencing at the *AR* gene on chromosome X (P50-H5). Blue indicates low methylation probability and red high methylation probability. In this sample, targeted read-out was suggestive for a 20-80% skewing of the X-chromosome, which cannot be observed in the LRS methylation profile. **B** In contrast, for another sample (P50-E8), the 100% skewing can be observed at the *AR* gene. For each sample, the panels show phased alleles, denoted as hap1 and hap2. Yellow arrow indicates the locus where X-(in)activation is measured when targeted testing is employed.

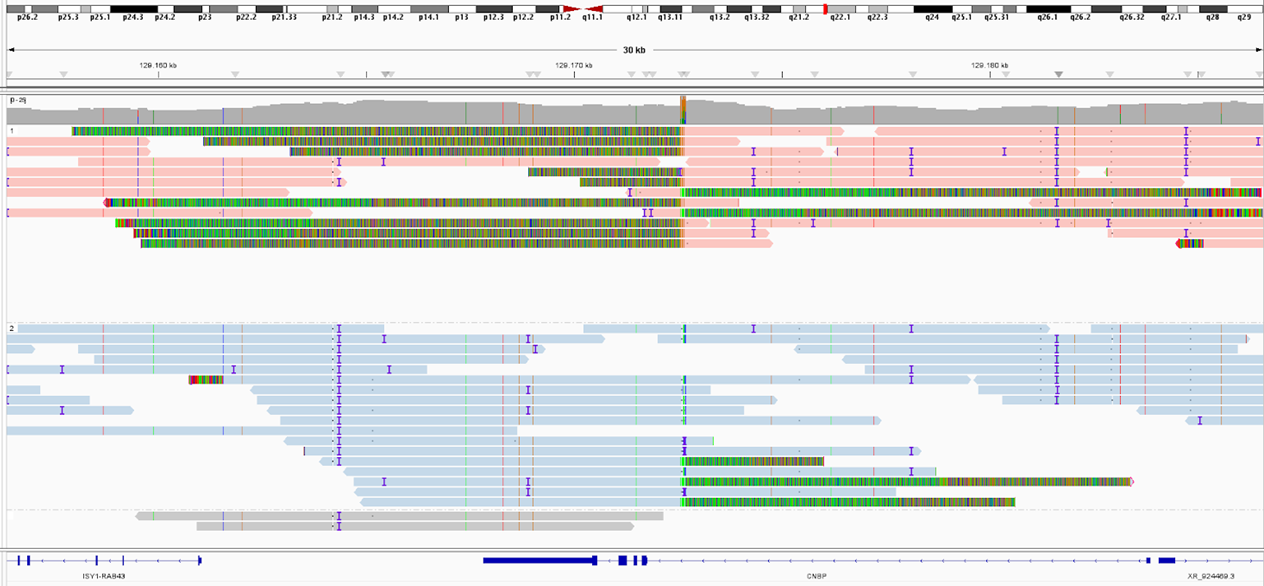

**Figure S2-3: *CNBP* repeat expansion.**  IGV Screenshot of the *CNBP* locus with reads colored by allele for sample P50-H1. Red reads show clear indicators of repeat expansions in the upper panel with clipped sequences and increased coverage. Analysis of this locus with TRGT failed because the (sequence specific) repeat motif was not part of the TRGT configuration. Manual addition of the motif to this TRGT configuration subsequently identified the repeat expansion automatically.

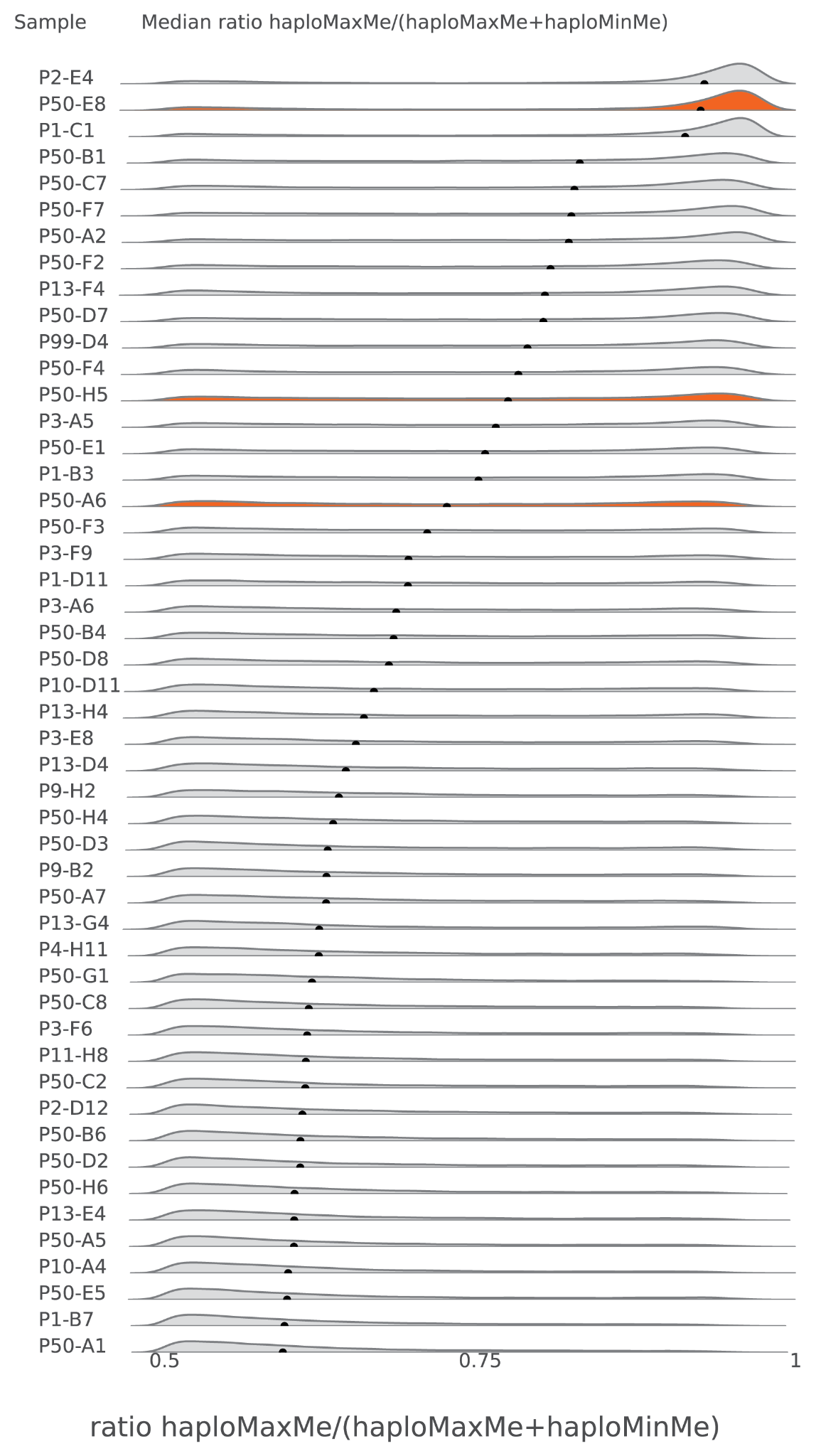

**Figure S3**. Basic assessment of skewed X-inactivation for all female samples for the X chromosome (**Methods**). Three samples show a clear skewing: P2-E4 (with a translocation on chrX), P50-E8 (mother of a son with X-linked disorder) and P1-C1 (with a duplication on chrX). Other samples showed less obvious skewing. The samples originally suspected to be X-inactivation skewed are highlighted in orange (P50-E8, P50-H5, P50-A6).

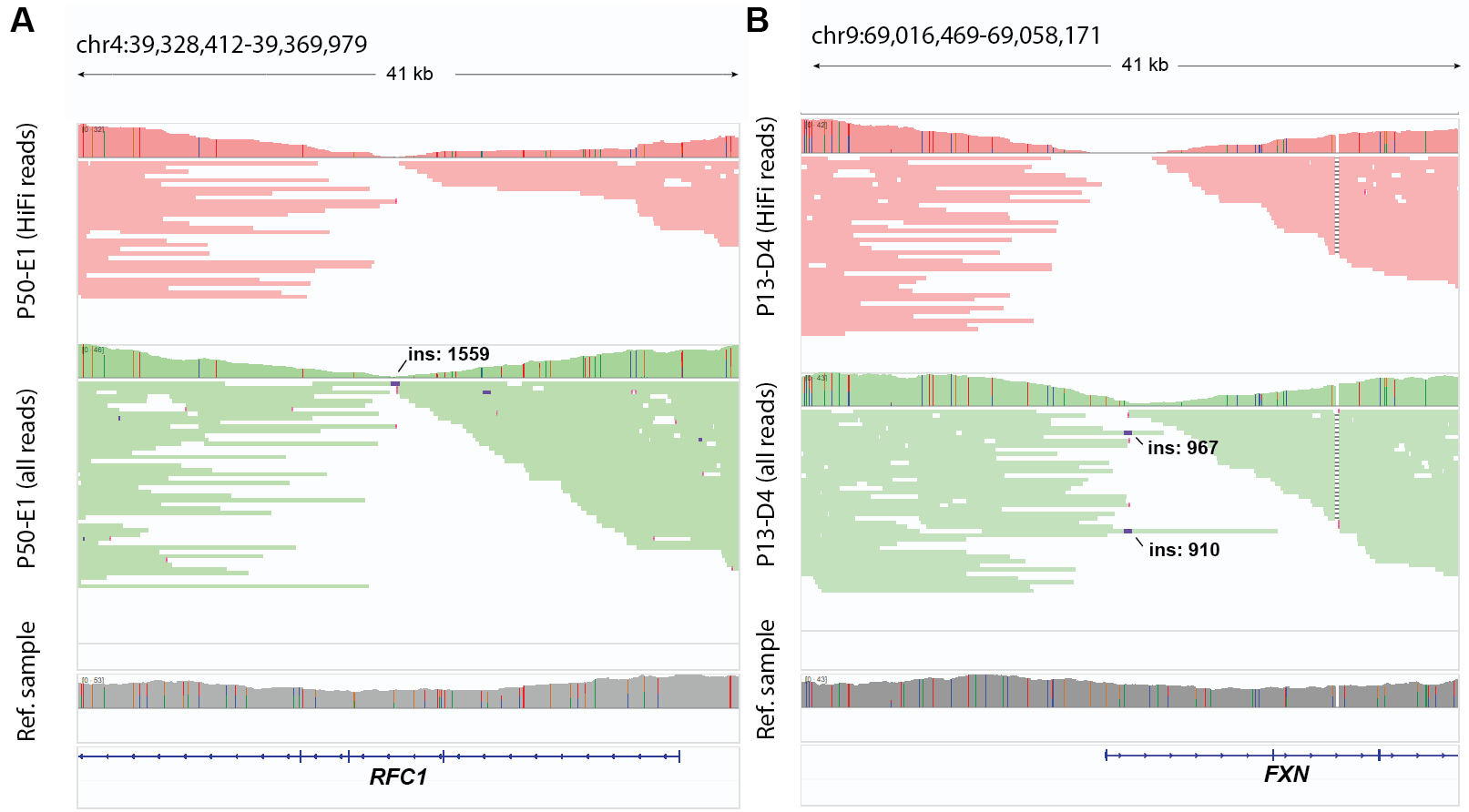

**Figure S4**. IGV screenshot of the attempted rescue of coverage of AG-rich regions by including rejected reads. **(A)** Top panel shows the original data for sample P50-E1 based on HiFi reads (red) where there is no sequence coverage at the *RFC1* locus. Middle panel shows that adding additional non-HiFi reads (green) for the *RFC1* locus did not lead to sufficient additional reads with the repeat expansion to identify the expansion by visual inspection. Bottom panel shows that the coverage profile of a reference sample without a repeat expansion is not aberrant at this site. **(B)** Top panel shows the original data for sample P13-D4 based on HiFi reads (red) where there is no sequence coverage at the *FXN* locus. Middle panel shows the locus after inclusion of non-HiFi reads (green) with sufficient coverage to manually identify the repeat expansion. Bottom panel shows that the coverage profile of a reference sample without a repeat expansion is not aberrant at this site.

**Table S1.** Overview of 145 variants from 100 patient samples selected for this study

External File: Table S1.xlsx

Columns left to right: Patient Index Number, Variant Identifier, Patient Identifier, Patient Gender, Reason for inclusion in the study, Single- or Multi- gene variant, Gene locus, Variant details, Variant zygosity (heterozygous/homozygous), Genetic tests employed for initial variant discovery, Variant class, Variant type, Method of detection in HiFi genome (automated, visual or not at all). The last columns indicate whether a particular variant was re-called by different LRS based callers: *deep variant* (SNVs, Indels), *TRGT* (short tandem repeats; STRs), *paraphase* (variants in duplicated regions / pseudogenes), hifiCNV (copy number variants; CNVs), pbsv (Structural variants; SVs).

**Table S2.** Quality control of 100 samples PacBio HiFi samples

External File: Table S2.xlsx

Columns from left to right: Patient identifier, Total number of HiFi reads, Total HiFi sequencing yield (Gb), average genome sequence coverage, average HiFi read length (bp), and HiFi average Q value.

**Table S3**. Repeat expansions with high AG-content missed by LRS, after attempted rescue with rejected reads. Columns indicate (from left to right): sample identifier, STR locus by gene name, expected repeat expansion, sequence coverage with HiFi reads at the locus, sequencing reads observed by visual inspection in IGV, sequence coverage with HiFi and non-HiFi reads, sequencing reads observed by visual inspection in IGV when including non-HiFi reads.

| **Sample** | **STR** | **Motif** | **Expected** | **Coverage** | **Observed** | **Coverage with rejected reads** | **Observed with rejected reads** |
| --- | --- | --- | --- | --- | --- | --- | --- |
| P13-D4 | FXN | GAA | (GAA)>65 (homozygous) | 0 | no result | 13.95 | GAA (309/328) |
| P50-E1 | RFC1 | AAGGG | (AAGGG)1500 / (AAGGG)1200 | 0.071 | no results | 1.16 | AAGGG (244/244), GGGAC (77/77), AAAGGG (5/5) |
| P50-F1 | RFC1 | AAGGG | (AAGGG)1200 / (AAAAG)11 | 10.1 | AAAAG(10/10) | 18.39 | AAAAG(10/10) |
| P50-G1 | RFC1 | AAGGG | (AAGGG)770 / (ACAGG)1500 | 6.82 | AAAAG(124/125), AAAGG (93,272), AGAGG(763/312), GGGAC(512/999) | 24.14 | AAAAG(124/123), AAAGG (266/548), GGGAC(1050/876) |

**Table S4**. Detailed results of the downsampling experiment

External File: Table S4.xlsx

Sheet 1: Details of the variants tested for re-call in the downsampling experiments. Columns left to right: Patient Identifier, Variant Identifier, Variant Caller (snv: DeepVariant; para/para_json: Paraphase, pbsv: pbsv, hifiasm: hifiasm, str:TRGT), Variant type, Genomic location of variant call, Variant-specific info; e.g. substitution type, SV length, pseudogene copy numbers. Sheet 2: Recovery statistics of variants, stratified by variant source and coverage. Columns left to right: Target downsampling coverage, Source of variant call, Median percent of re-called variants compared to full dataset (30X). Sheet 3: Full list of all variants tested across all samples, with indication for successful or failed recall (‘found’; last column) Remaining columns: similar to Sheet 1.

**Table S5**. Cross comparison of the 42 samples tested by both short and long read sequencing.

External File: Table S5.xlsx

Cross comparison of 42 samples with 70 variants tested by both short read genome sequencing (30x) and long read genome sequencing (30x). HiFi genomes followed by standardized variant callers increase the number of automated detection from 28/70 (41%) to 62/70 (89%). Concomitantly, the variants that remain undetected are reduced from 20/70 (29%) in short reads to 5/70 (7%) in long read genomes.

**Table S6.** Details of the 42 samples tested by both short and long read sequencing.

External File: Table S6.xlsx

Details of the 42 samples with 70 variants tested by both short read genome sequencing (30x) and long read genome sequencing (30x). Columns left to right: Patient Index Number, Variant Identifier, Patient Identifier, Reason for inclusion in the study, Single- or Multi- gene variant, Gene locus, Variant details, Variant type, Method of detection in HiFi genome (automated, visual or not at all), Detection in SR-WGS in Schobers et al. (automated, visual or not at all).

**Supplemental methods**

**Cohort selection**

We retrospectively selected archival residual DNA material from a cohort (n = 100) with known clinically relevant variants (n = 145) from genome diagnostic laboratories of the Radboudumc in Nijmegen and the MUMC + in Maastricht. The samples were selected based on pathogenic variants being located in a region difficult to detect using short read sequencing and/or require routine detection using a workflow difficult to replace by short read genome sequencing as previously determined by Schobers et al. 2024. In brief, this results in a series of samples with STR repeat expansions, samples with variants in homologous regions/pseudogenes, and complex events involving a myriad of different types of structural variation (e.g. translocations, variants >2 chromosomal breaks). These variants typically require a combination of different diagnostic workflows, such as karyotyping, triplet repeat-primed PCR and fragment analysis, exome sequencing and MLPA and/or sanger sequencing. Of note, 42 of 100 samples were previously tested in the technical benchmarking study (**Table S1**). Importantly, for the purpose of this study, each sample is numbered; yet, if a sample contains multiple clinically relevant variants, each variant is also individually numbered (**Table S2**). This resulted in 145 variants in 100 samples. Based on the selection criteria, the cohort is considered representative for our diagnostic centers when focussing on variants challenging to detect using short read sequencing technologies as first tier tests.

The study conforms to the principles of the Helsinki declaration. In addition, the study was performed as part of a local validation study for the implementation of GS under ISO15189 accreditation and assessed as a diagnostic innovation by the Medical Ethics Review Committee Arnhem-Nijmegen under dossier number 2020–7142.

**Long read genome sequencing and data analysis**

For the LRS, we targeted 30 × HiFi coverage by using 1 SMRT Cell per sample (**Table S2**). All samples were processed in the same fashion, according to the manufacturer’s instructions (PacBio, Menlo Park, CA, USA). In brief, 7 µg gDNA was sheared on Megaruptor 3 (Diagenode, Liège, Belgium) to a target size of 15-18 kb, libraries were prepared with SMRTbell prep kit 3.0 (PacBio, Menlo Park, CA, USA), size-selected > 10 kb on the BluePippin (Sage Science, Beverly, MA, USA), and sequenced for 24 h on the Revio system (ICS 12.0.4).

Samples were analysed using an analysis pipeline that incorporates different PacBio software tools for Long read sequencing (LRS) data analysis. Alignment (pbmm2 v1.10.0) of High Fidelity (HiFi) reads if performed against the GRCh38 reference genome, and generates haplotags by performing de novo assembly with Hifiasm (v.0.15.3). Structural variants (PBSV v2.9.0) and Small variants (DeepVariant v1.5.0) are called with phasing information (HiPhase v0.10.1) and annotated using publicly available databases. A specific analysis for Copy Number Variants is performed (HifiCNV v0.1.6) by detecting variation in depth of coverages. Short tandem repeats are called (TRGT v0.4.0), visualize (TRVZ v0.4.0) and annotated using an inhouse pipeline. Specific variant calling for some paralogs and pseudogenes are called using Paraphase v.2.2.3. Methylation calls were generated using pb-cpg-tools (v.2.3.1).

**Variant detection strategy**

For all samples relevant output files from software tools, depending on the variant class were checked for the detection of the variant, taking into account a suitable genomic window. When variants were not detected by the software the region of the variant was visually inspected using IGV version 2.17.0 , with the following settings. A variant was considered to be detected by visual inspection when there were clear aberrations visible in the sample at the relevant locus that were not present in other samples. This included clipped reads, read misalignments, abnormal sequence coverage, deviant variant patterns or aberrant methylation. Overall we considered whether based on the sequencing data it is plausible that the variant could have reasonably been detected.

**De-novo assembly.** Local de-novo assembly was performed by extracting reads mapping within a window of 5Mbp (in hg38) around the variant of interest. *Hifiasm* version 0.15.3 was invoked with default parameters, and assemblies were considered successful if the variant was entirely contained inside one unitig, i.e. phased in its entirety. Visualization of the resulting assembled sequences was done with *NAHRwhals* version 1.4.

**X-inactivation.** Haplotype-specific methylation data was generated using CpGTools of Pacbio. CpG sites for ascertaining chromosome X inactivation were selected based three criteria: intersecting a CpG island (UCSC CpG islands on chrX (Karolchik et al. 2004)), measured in 25 male samples, and finally having a mean methylation score < 5. This resulted in a total of 16,109 sites. Within these sites within the female samples, the haplotype methylation scores were categorized as haplotype Maximum Methylation (hapMax), or haplotype Minimum methylation (hapMin) based on their methylation score. The hapMax/(hapMax + hapMin) ratio was calculated for each sample and the resulting distributions were plotted in a ridgeplot using seaborn.

**Regions of homozygosity.** To visually investigate suspected regions of homozygosity, we extracted coordinates and VAF (variant allele frequency) labels of all variants called by DeepVariant. The resulting data track was converted to .bedpe format and visualized using IGV v2.16.2.

**Data titration.** *In-silico* downsampling experiments were implemented as follows. Aligned read files (.bam) from 97 out of 100 samples were subsampled to target coverages of 10X, 15X and 20X using the “*samtools view -b”* command (samtools version 1.11). This process was repeated 10 times for each target coverage, resulting in a total of 30 subsampled readsets per sample, or 10 per target coverage. Downstream processing and variant calling was done analogously to the original datasets, resulting in independent variant callsets for each permutation. Three samples (P13-G4, P50-G5, P50-H7) with an initial coverage of less than 20-fold were excluded from this analysis.

**Titration analysis.** 97/100 samples had a coverage of at least 20-fold (median 29.8x) and were thus considered for our titration analysis. In the original dataset, these samples together carried 117 pathogenic biological variants which could be automatically detected. Due to cross-calling between different tools (e.g., hifiCNV and pbsv both calling certain CNVs), those 117 variants were represented by 177 individual variant calls. We checked for re-call of these 177 calls in the subsampled datasets with a script implementing the following criteria ([**https://github.com/WHops/lrs100_downsample**](https://github.com/WHops/lrs100_downsample)).

- **SNPs**: Same substitution within ±1bp.
- **Mitochondrial SNPs: additionally**: Affected fraction within 20 percent points of original call.
- **Indels (<50 bp)**: All breakpoints within ±10 bp, and event length within ±5 bp of the original call.
- **SVs (>50 bp, excl. translocations)**: The re-call and original call share ≥50% reciprocal overlap.
- **Translocations**: All breakpoints found within ±50bp of the original call.
- **STRs**: Within ±1kbp: TRGT motif copy number on both alleles identified (±20% CN deviation allowed).
- **Gene/Pseudogene CNVs (Paraphase):** The total copy number of genes + their pseudogenes is reported correctly.

HifiCNV sometimes reports ‘split’ calls, i.e. multiple consecutive calls that cover a single actual event. To alleviate this potential discrepancy, both the original calls and the permuted calls are always first merged with distance up to 1000 bp, joining adjacent segments.

For STRs, we test for the motif and copy-number of both alleles and allow for plus/minus 20% in both. In cases where the copy number of the motif is denoted as ‘>n copy numbers’, we test for that allele only that there is at least this amount of copy numbers.

Source ‘para_json’ denotes variants that are exclusively called in the paraphrase json files. Those are copy number changes affecting Gene/Pseudogene combinations. There exists an edge case if e.g. a copy-number change is correctly noted, but ascribed to the wrong gene copy. In such cases, we accept that paraphrase has ‘in principle’ identified the variant and count this variant as recovered.
